# Mendelian randomization analysis identified genes potentially pleiotropically associated with periodontitis

**DOI:** 10.1101/2021.02.05.21251192

**Authors:** Feng Wang, Di Liu, Yong Zhuang, Bowen Feng, Wenjin Lu, Jingyun Yang, Guanghui Zhuang

**Author notes:** Correspondence to Guanghui Zhuang, Department of Stomatology, The First Affiliated Hospital of Dalian Medical University, 222 Zhongshan Rd, Xigang District, Dalian, Liaoning 116011, China, or Jingyun Yang, Rush Alzheimer’s Disease Center, Rush University Medical Center, 1750 West Harrison Street, Suite 1000, Chicago, IL 60612, USA.

## Abstract

**Objective:** To prioritize genes that were pleiotropically or potentially causally associated with periodontitis.

**Methods:** We applied the summary data-based Mendelian randomization (SMR) method integrating genome-wide association study (GWAS) for periodontitis and expression quantitative trait loci (eQTL) data to identify genes that were pleiotropically associated with periodontitis. We performed separate SMR analysis using CAGE eQTL data and GTEx eQTL data. SMR analysis were done for participants of European and East Asian ancestries, separately.

**Results:** We identified multiple genes showing pleiotropic association with periodontitis in participants of European ancestry and participants of East Asian ancestry. *PDCD2* (corresponding probe: ILMN_1758915) was the top hit showing pleotropic association with periodontitis in participants of European ancestry, and *BX093763* (corresponding probe: ILMN_1899903) and AC104135.3 (corresponding probe: ENSG00000204792.2) were the top hits in participants of East Asian ancestry using CAGE eQTL data and GTEx eQTL data, respectively.

**Conclusion:** We identified multiple genes that may be involved in the pathogenesis of periodontitis in participants of European ancestry and participants of East Asian ancestry. Our findings provided important leads to a better understanding of the mechanisms underlying periodontitis and revealed potential therapeutic targets for the effective treatment of periodontitis.

## Introduction

Periodontitis is a common disease characterized by an inflammatory response to commensal and pathogenic oral bacteria^1^. The primary clinical features of periodontitis include periodontal pocketing, alveolar bone loss (BL), clinical attachment loss (CAL), and gingival inflammation^2^. Based on the 2009lJ2014 National Health and Nutrition Examination Surveys data, it was estimated that periodontitis affected about 42% of US adults aged 30 to 79 years^3^. Periodontitis is considered as the main cause of tooth loss in adults. Moreover, it is also associated with various systemic conditions such as coronary heart disease^4^, diabetes^5^ and pre-term birth^6^. Periodontitis not only affects a patient’s life, it also brings tremendous economic burden to the society, with an estimated global productivity loss due to untreated severe periodontitis being around $38.85 billion in 2015^7^.

Periodontitis is a complex, multi-factorial infectious disease with possible contributions from multiple factors, including immunological response^8^, oral bacterial infections^9^, lifestyle factors such as smoking^10^ and alcohol consumption^11^, psychological factors such as stress^12^ and depression^13^, and systematic diseases such as diabetes^14^. Previous studies also suggested that genetics plays an important role in the pathogenesis of periodontitis. For example, genetically identical monozygotic twins have more than a twofold increased risk of early onset periodontitis, compared with dizygotic twins^15^. Another population-based twin study estimated that the heritability of periodontitis was approximately 50%^16^. Moreover, many GWAS and candidate gene studies have identified a number of genetic loci associated with the susceptibility of periodontitis^17-22^. However, the biological mechanisms of these findings remain largely unclear, and more studies are needed to explore genes that are potentially causally associated with periodontitis to better understand the pathogenesis of periodontitis.

Mendelian randomization (MR) uses genetic variants as the proxy to randomization and is a promising tool to search for pleotropic/potentially causal effect of an exposure (e.g., gene expression) on the outcome (e.g., periodontitis) without the need of conducting conventional randomized clinical trials (RCTs)^23^. Confounding and reverse causation, which are commonly encountered in traditional association studies, can be greatly reduced by using MR. MR has been successful in identifying gene expression probes or DNA methylation loci that are pleiotropically/potentially causally associated with various phenotypes, such as neuropathologies of Alzheimer’s disease and severity of COVID-19^24,25^.

In this study, we applied the summary data-based MR (SMR) method integrating summarized GWAS data for periodontitis and cis-eQTL (expression quantitative trait loci) data to prioritize genes that are pleiotropically/potentially causally associated with periodontitis.

## Methods

### Data sources

#### eQTL data

In the SMR analysis, cis-eQTL genetic variants were used as the instrumental variables (IVs) for gene expression. We performed SMR analysis using gene expression data in blood due to the unavailability of eQTL data in the eye. Specifically, we used the CAGE eQTL summarized data^26^, which included 2,765 participants, and the V7 release of the GTEx eQTL summarized data^27^, which included 338 participants. The eQTL data can be downloaded at https://cnsgenomics.com/data/SMR/#eQTLsummarydata.

#### GWAS data for periodontitis

The GWAS summarized data were provided by a recent genome-wide association meta-analysis of periodontitis^28^. The results were based on meta-analyses of 1000 genomes phase 1 version 2/3 imputed GWASs on periodontitis, with a total of nine cohorts from the Gene-Lifestyle Interactions in Dental Endpoints (GLIDE) consortium^29^. Specifically, the meta-analysis for participants of European ancestry included seven cohorts with a total sample size of 45,563 (17,353 cases and 28,210 controls), and the meta-analysis for participants of East Asian ancestry included two cohorts with a total sample size of 17,350 (1,680 cases and 15,670 controls). All participating studies assumed an additive genetic model, adjusting for age, age-squared and other study-specific covariates. The GWAS summarized data can be downloaded at https://data.bris.ac.uk/data/dataset/2j2rqgzedxlq02oqbb4vmycnc2.

#### SMR analysis

We conducted the SMR analysis with cis-eQTL as the IV, gene expression as the exposure, and periodontitis as the outcome. The analysis was done using the method as implemented in the software SMR. Detailed information regarding the SMR method was reported in a previous publication^30^. In brief, SMR applies the principles of MR to jointly analyze GWAS and eQTL summary statistics in order to test for pleotropic association between gene expression and a trait due to a shared and potentially causal variant at a locus. We also conducted the heterogeneity in dependent instruments (HEIDI) test to evaluate the existence of linkage in the observed association. A P_HEIDI_ of less than 0.05 indicates that the observed association could be due to two distinct genetic variants in high linkage disequilibrium with each other. We adopted the default settings in SMR (e.g., minor allele frequency [MAF] > 0.01, removing SNPs in very strong linkage disequilibrium [LD, r^2^ > 0.9] with the top associated eQTL, and removing SNPs in low LD or not in LD [r^2^ <0.05] with the top associated eQTL) except relaxing the threshold of eQTL P-value (P_eQTL_ <10^−4^) due to the exploratory nature of this study, and used false discovery rate (FDR) to adjust for multiple testing. We performed SMR analysis for participants of European and East Asian ancestries, separately, using CAGE and GTEx eQTL data, respectively, comprising a total of four SMR analyses.

We used Affymetrix exon array S1.0 platforms to annotate the transcripts. We conducted functional enrichment analysis using the functional annotation tool “Metascape” for the top tagged genes to functionally annotate putative transcripts^31^. Gene symbols corresponding to the ten top hit genes were used as the input of the gene ontology (GO) and Kyoto Encyclopedia of Genes and Genomes (KEGG) enrichment analysis.

Data cleaning and statistical/bioinformatical analysis was performed using R version 4.0.3 (https://www.r-project.org/), PLINK 1.9 (https://www.cog-genomics.org/plink/1.9/) and SMR (https://cnsgenomics.com/software/smr/).

## Results

### Basic information of the summarized data

The number of participants of the CAGE eQTL data is much larger than that of the GTEx eQTL data, so is the number of eligible probes. The sample size of the GWAS data for the European ancestry is much large than that for the East Asian ancestry, so is the number of eligible genetic variants. The detailed information was shown in **Table 1**.

**Table 1.** Basic information of the GWAS and eQTL data.

| **Data Source** | **Total number of participants** | **Number of eligible genetic variants or probes** |
| --- | --- | --- |
| **European ancestry** |  |  |
| **eQTL data** |  |  |
| **CAGE** | 2,765 | 8,230 |
| **GTE<sub>x</sub>** | 338 | 2,162 |
| **GWAS data** | 45,563 | 779,1334 |
| **East Asian ancestry** |  |  |
| **eQTL data** |  |  |
| **CAGE** | 2,765 | 7,304 |
| **GTE<sub>x</sub>** | 338 | 2,010 |
| **GWAS data** | 17,350 | 418,8352 |

GWAS: genome-wide association studies; QTL, quantitative trait loci

### SMR analysis in participants of European ancestry

In participants of European ancestry, we identified two genes showing pleiotropic association with periodontitis after correction for multiple testing using FDR (**Table 2**). Specifically, using the CAGE eQTL data, our SMR analysis identified two genes that were pleiotropically/potentially causally associated with periodontitis, including *PDCD2* (ILMN_1758915; P_SMR_=3.77×10^−5^; **Figure 1**) and *D4S234E* (i.e., *NSG1*, ILMN_1772627; P_SMR_=9.08×10^−4^; **Figure 2**). GO enrichment analysis of biological process and molecular function showed that the significant genes were involved in two GO terms, including positive regulation of cysteine-type endopeptidase activity (GO:2001056) and positive regulation of defense response (GO:0031349; **Figure S1A**). Concept network analysis of the ten top hit genes also revealed multiple domains related with endopeptidase activity (**Figure S1B**). More information could be found in **Table S1**.

**Table 2.** The top ten probes identified in the SMR analysis in participants of European ancestry.

| eQTL data | Probe ID | Gene | CHR | Top SNP | P<sub>eQTL</sub> | P<sub>GWAS</sub> | Beta | SE | P<sub>SMR</sub> | P<sub>HEIDI</sub> | N<sub>SNP</sub> |
| --- | --- | --- | --- | --- | --- | --- | --- | --- | --- | --- | --- |
| **CAGE** | ILMN_1758915 | *PDCD2* | 6 | rs17875294 | $7.50 \times 10^{-74}$ | $2.34 \times 10^{-5}$ | 0.15 | 0.04 | **$3.77 \times 10^{-5}$** | $4.52 \times 10^{-1}$ | 20 |
| | ILMN_1772627 | *NSG1* | 4 | rs6843595 | $7.10 \times 10^{-291}$ | $8.45 \times 10^{-4}$ | 0.05 | 0.02 | **$9.08 \times 10^{-4}$** | $1.33 \times 10^{-1}$ | 20 |
| | ILMN_1823130 | *F01764* | 4 | rs2369111 | $1.74 \times 10^{-17}$ | $3.62 \times 10^{-4}$ | 0.24 | 0.07 | $1.00 \times 10^{-3}$ | $3.31 \times 10^{-2}$ | 20 |
| | ILMN_1808251 | *C9orf38* | 9 | rs4556138 | $6.67 \times 10^{-10}$ | $1.94 \times 10^{-4}$ | 0.35 | 0.11 | $1.40 \times 10^{-3}$ | $1.51 \times 10^{-2}$ | 20 |
| | ILMN_1748915 | *S100A12* | 1 | rs3014878 | $2.17 \times 10^{-177}$ | $1.45 \times 10^{-3}$ | -0.06 | 0.02 | $1.58 \times 10^{-3}$ | $3.77 \times 10^{-1}$ | 20 |
| | ILMN_1748221 | *PADI6* | 1 | rs1535876 | $2.45 \times 10^{-50}$ | $1.69 \times 10^{-3}$ | -0.12 | 0.04 | $2.13 \times 10^{-3}$ | $3.50 \times 10^{-1}$ | 20 |
| | ILMN_1659511 | *LOC645652* | 1 | rs10927894 | $3.79 \times 10^{-7}$ | $1.38 \times 10^{-4}$ | 0.60 | 0.20 | $2.30 \times 10^{-3}$ | $2.88 \times 10^{-2}$ | 20 |
| | ILMN_1710937 | *IFI16* | 1 | rs12122315 | $5.38 \times 10^{-19}$ | $1.27 \times 10^{-3}$ | -0.22 | 0.07 | $2.46 \times 10^{-3}$ | $1.37 \times 10^{-1}$ | 20 |
| | ILMN_1729801 | *S100A8* | 1 | rs58644524 | $1.93 \times 10^{-19}$ | $1.36 \times 10^{-3}$ | -0.20 | 0.06 | $2.52 \times 10^{-3}$ | $3.72 \times 10^{-1}$ | 20 |
| | ILMN_2096405 | *WDR37* | 10 | rs12768746 | $3.40 \times 10^{-7}$ | $2.40 \times 10^{-4}$ | 0.41 | 0.14 | $2.91 \times 10^{-3}$ | $4.89 \times 10^{-2}$ | 20 |
| **GTE<sub>x</sub>** | ENSG00000168824.10 | *NSG1* | 4 | rs6414635 | $1.94 \times 10^{-41}$ | $9.57 \times 10^{-4}$ | 0.08 | 0.02 | $1.35 \times 10^{-3}$ | $2.34 \times 10^{-1}$ | 20 |
| | ENSG00000256049.2 | *PADI6* | 1 | rs10888031 | $2.95 \times 10^{-42}$ | $1.32 \times 10^{-3}$ | -0.06 | 0.02 | $1.74 \times 10^{-3}$ | $3.22 \times 10^{-1}$ | 20 |
| | ENSG00000163221.7 | *S100A12* | 1 | rs57572338 | $1.56 \times 10^{-15}$ | $7.58 \times 10^{-4}$ | -0.37 | 0.12 | $1.90 \times 10^{-3}$ | $5.90 \times 10^{-1}$ | 9 |
| | ENSG00000184985.12 | *SORCS2* | 4 | rs62289059 | $8.07 \times 10^{-15}$ | $8.95 \times 10^{-4}$ | 0.15 | 0.05 | $2.23 \times 10^{-3}$ | $4.30 \times 10^{-1}$ | 20 |
| | ENSG00000127952.12 | *STYXL1* | 7 | rs115332207 | $3.41 \times 10^{-39}$ | $2.22 \times 10^{-3}$ | 0.12 | 0.04 | $2.94 \times 10^{-3}$ | $9.10 \times 10^{-1}$ | 20 |
| | ENSG00000233609.3 | *RP11-62H7.2* | 8 | rs13259143 | $5.63 \times 10^{-19}$ | $2.60 \times 10^{-3}$ | 0.16 | 0.06 | $4.40 \times 10^{-3}$ | $6.75 \times 10^{-2}$ | 20 |
| | ENSG00000106804.6 | *C5* | 9 | rs7036980 | $2.84 \times 10^{-10}$ | $1.63 \times 10^{-3}$ | 0.19 | 0.07 | $4.76 \times 10^{-3}$ | $6.56 \times 10^{-1}$ | 20 |
| | ENSG00000213523.5 | *SRA1* | 5 | rs76128141 | $7.51 \times 10^{-13}$ | $2.88 \times 10^{-3}$ | -0.27 | 0.10 | $5.91 \times 10^{-3}$ | $4.22 \times 10^{-2}$ | 14 |
| | ENSG00000163421.4 | *PROK2* | 3 | rs6777956 | $3.52 \times 10^{-17}$ | $4.01 \times 10^{-3}$ | -0.21 | 0.08 | $6.40 \times 10^{-3}$ | $5.76 \times 10^{-1}$ | 17 |
| | ENSG00000138835.18 | *RGS3* | 9 | rs41306506 | $6.49 \times 10^{-20}$ | $5.77 \times 10^{-3}$ | 0.20 | 0.08 | $8.22 \times 10^{-3}$ | $9.88 \times 10^{-1}$ | 20 |

\*The GWAS summarized data were provided by the study of Shungin et al. and can be downloaded at https://data.bris.ac.uk/data/dataset/2j2rqgzexlq02oqbb4vmcnc2. The CAGE and GTEx eQTL data can be downloaded at https://cnsgenomics.com/data/SMR/#eQTLsummarydata.
$P_{\text{eQTL}}$ is the P-value of the top associated cis-eQTL in the eQTL analysis, and $P_{\text{GWAS}}$ is the P-value for the top associated cis-eQTL in the GWAS analysis. Beta is the estimated effect size in SMR analysis, SE is the corresponding standard error, $P_{\text{SMR}}$ is the P-value for SMR analysis, $P_{\text{HEIDI}}$ is the P-value for the HEIDI test and N<sub>snp</sub> is the number of SNPs involved in the HEIDI test.
FDR was calculated at $P=10^{-3}$ threshold.
Bold font means statistical significance after correction for multiple testing using FDR.
CHR, chromosome; HEIDI, heterogeneity in dependent instruments; SNP, single-nucleotide polymorphism; SMR, summary data-based Mendelian randomization; QTL, quantitative trait loci; FDR, false discovery rate; GWAS, genome-wide association studies

**Figure 1.**
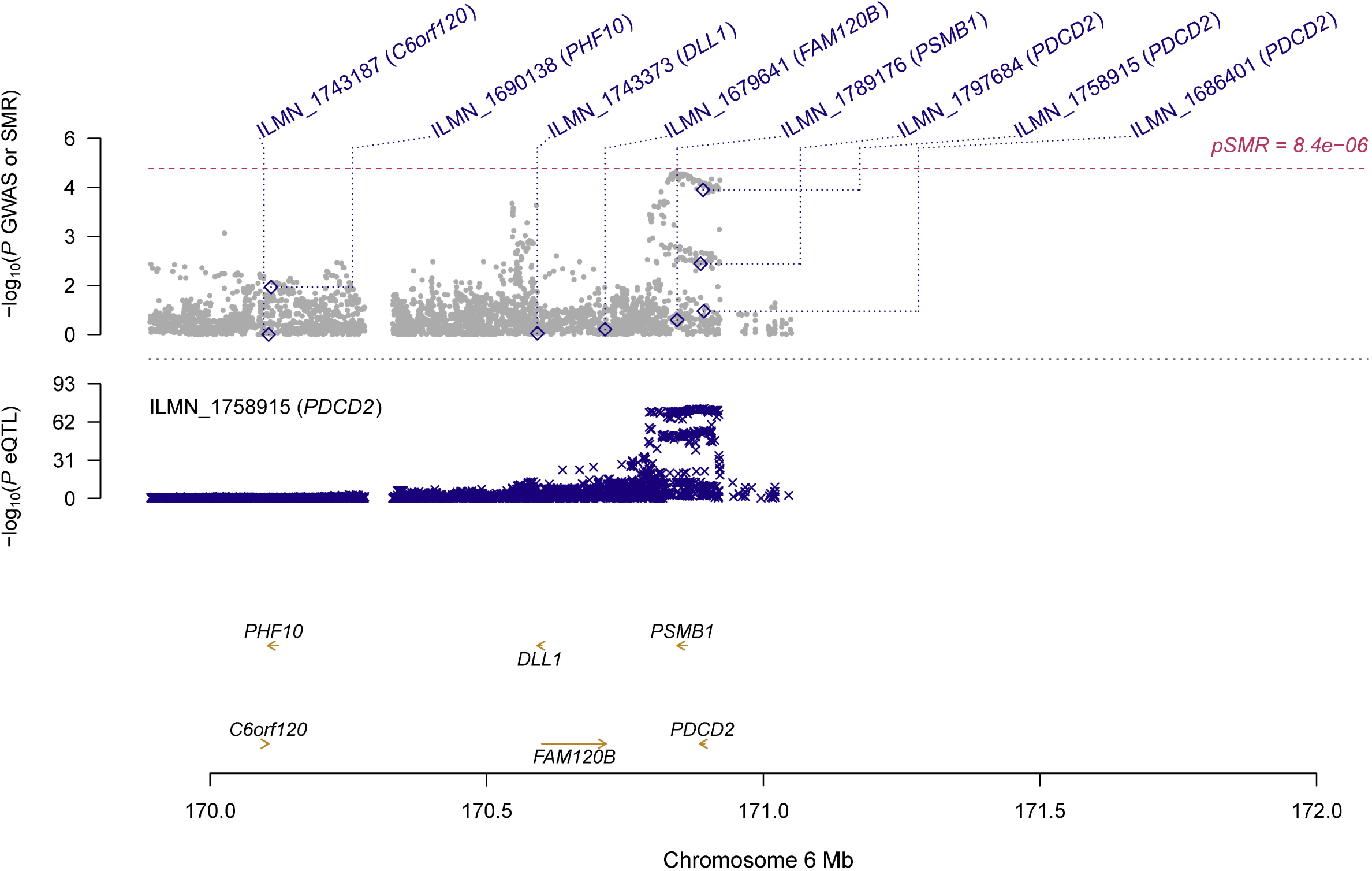
Prioritizing gene around *PDCD2* in association with periodontitis in participants of European ancestry. Top plot, grey dots represent the -log_10_(*P* values) for SNPs from the GWAS of periodontitis, and rhombuses represent the -log_10_(*P* values) for probes from the SMR test with solid rhombuses indicating that the probes pass HEIDI test and hollow rhombuses indicating that the probes do not pass the HEIDI test. Middle plot, eQTL results for ILMN_1758915 probe, tagging *PDCD2*. Bottom plot, location of genes tagged by the probes. GWAS, genome-wide association studies; SMR, summary data-based Mendelian randomization; HEIDI, heterogeneity in dependent instruments; eQTL, expression quantitative trait loci

**Figure 2.**
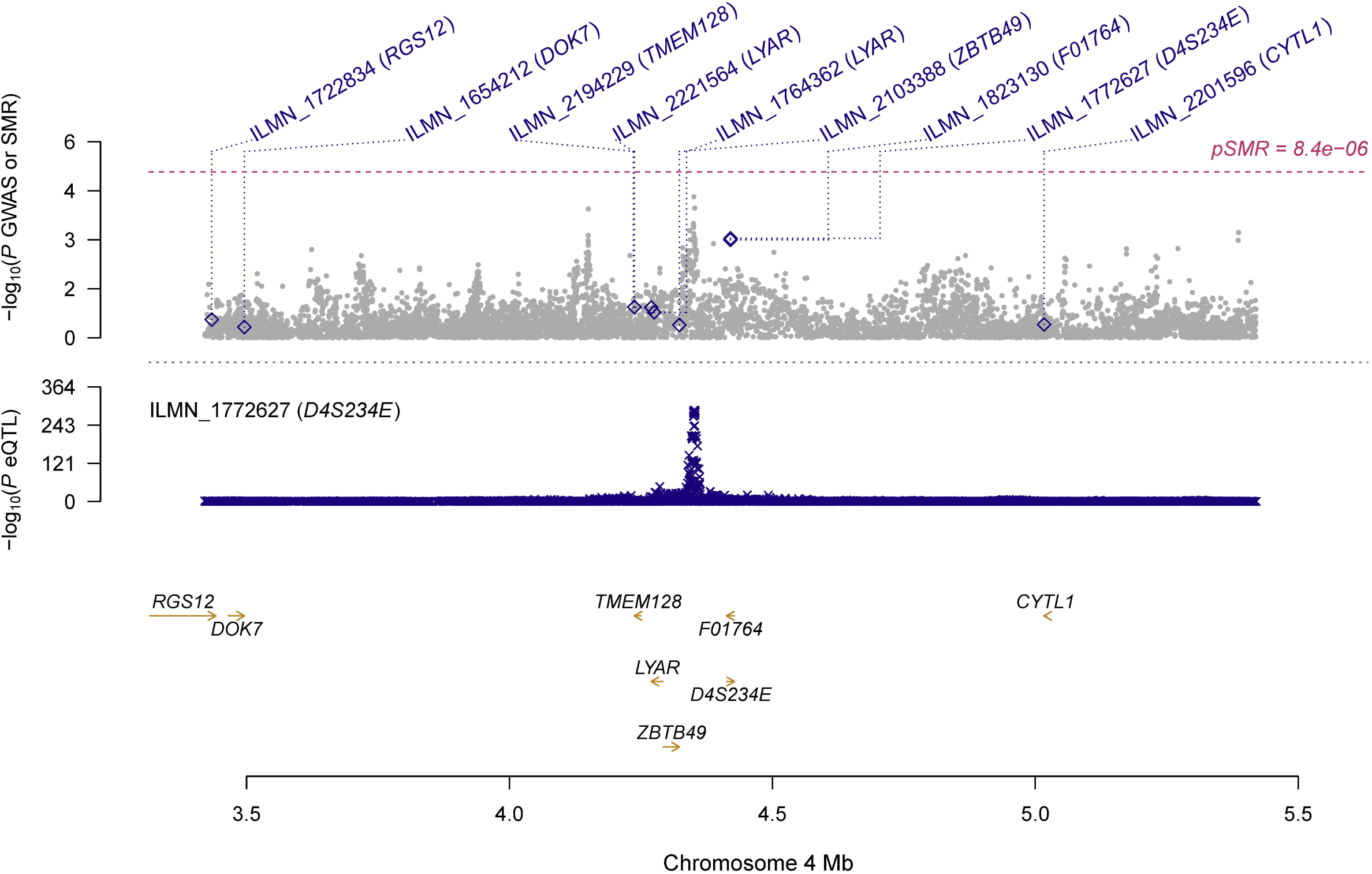
Prioritizing gene around *D4S234E* (i.e., *NSG1*) in association with periodontitis in participants of European ancestry. Top plot, grey dots represent the -log_10_(*P* values) for SNPs from the GWAS of periodontitis, and rhombuses represent the -log_10_(*P* values) for probes from the SMR test with solid rhombuses indicating that the probes pass HEIDI test and hollow rhombuses indicating that the probes do not pass the HEIDI test. Middle plot, eQTL results for ILMN_1772627 probe, tagging *D4S234E* (i.e., *NSG1*). Bottom plot, location of genes tagged by the probes. GWAS, genome-wide association studies; SMR, summary data-based Mendelian randomization; HEIDI, heterogeneity in dependent instruments; eQTL, expression quantitative trait loci

Using the GTEx eQTL data, we did not identify any genes that were pleiotropically/potentially causally associated with periodontitis, after correction for multiple testing using FDR (**Table 2**). However, we found that two genes, *NSG1* (CAGE eQTL: ILMN_1772627, P_SMR_=9.08×10^−4^; GETx eQTL: ENSG00000168824.10, P_SMR_=1.35×10^−3^) and *S100A12* (CAGE eQTL: ILMN_1748915, P_SMR_=1.58×10^−3^; GETx eQTL: ENSG00000163221.7, P_SMR_=1.90×10^−3^) were among the top hits in both SMR analyses. GO enrichment analysis of biological process and molecular function showed that the ten top hit genes were involved in two MAP kinase-related GO terms (GO:0043405 and GO:0043406; **Figure S1C**). Concept network analysis of the genes revealed multiple domains related with MAP kinase activity and inflammation (**Figure S1D**). More information could be found in **Table S2**.

### SMR analysis in participants of East Asian ancestry

In participants of East Asian ancestry, we identified two genes showing significant pleiotropic association with periodontitis after correction for multiple testing using FDR (**Table 3**). Specifically, using the CAGE eQTL data, our SMR analysis identified one gene, *BX093763* (ILMN_1899903, P_SMR_=2.33×10^−4^). GO enrichment analysis of biological process and molecular function showed that the ten top hit genes were involved in one GO terms, axon development (GO:0061564; **Figure S2A**). Concept network analysis of the genes revealed multiple domains related with inflammation (**Figure S2B**). More information could be found in **Table S3**. Using the GTEx eQTL data, our SMR analysis identified one gene, *AC104135*.*3*, that was pleiotropically/potentially causally associated with periodontitis, after correction for multiple testing using FDR (ENSG00000204792.2, P_SMR_=7.46×10^−4^; **Figure 3**). GO enrichment analysis of biological process and molecular function did not find any significant GO terms. Concept network analysis of the genes revealed multiple domains related with endogenous peptide antigen (**Figure S2C**). More information could be found in **Table S4**.

**Table 3.**
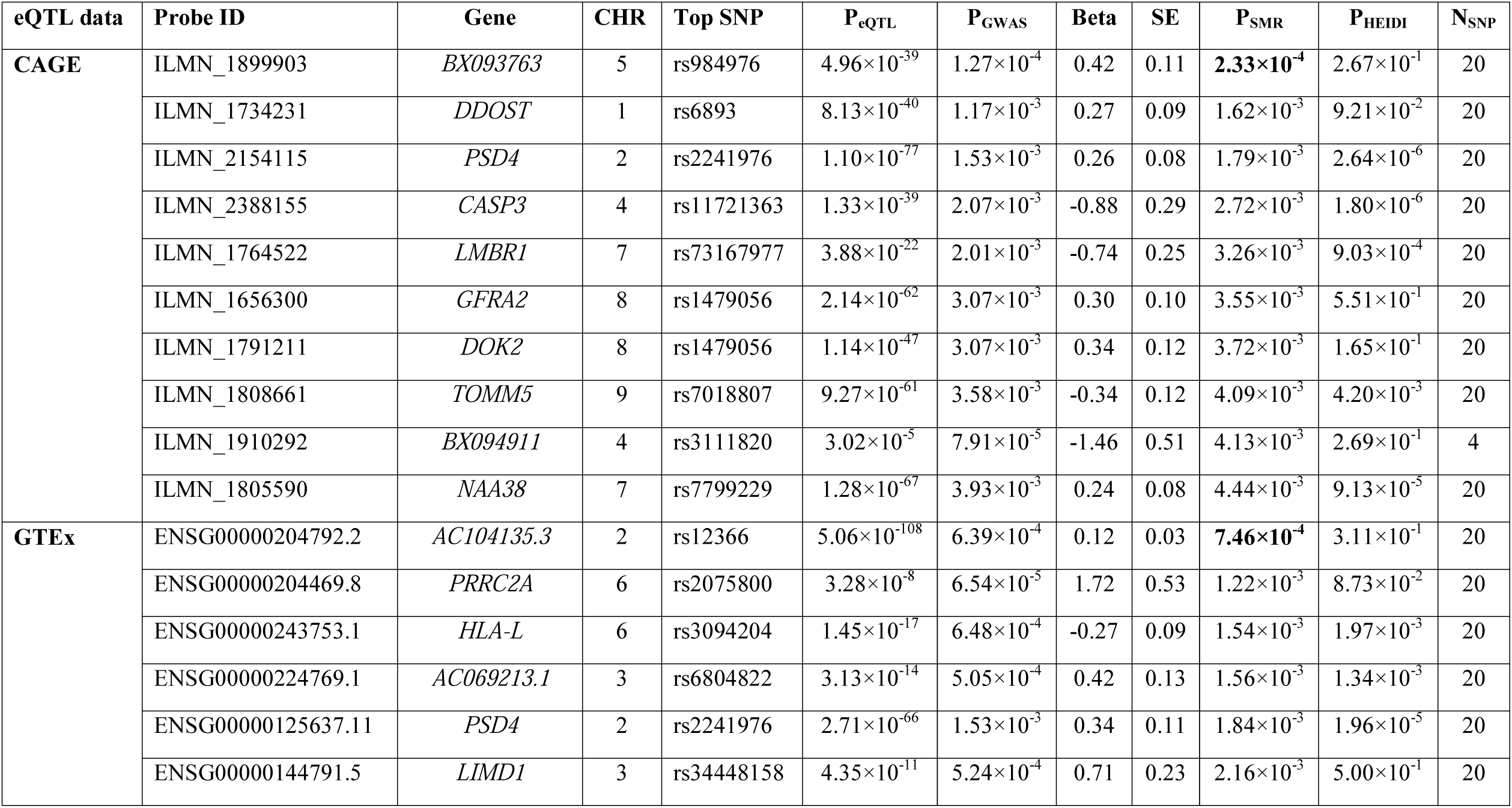
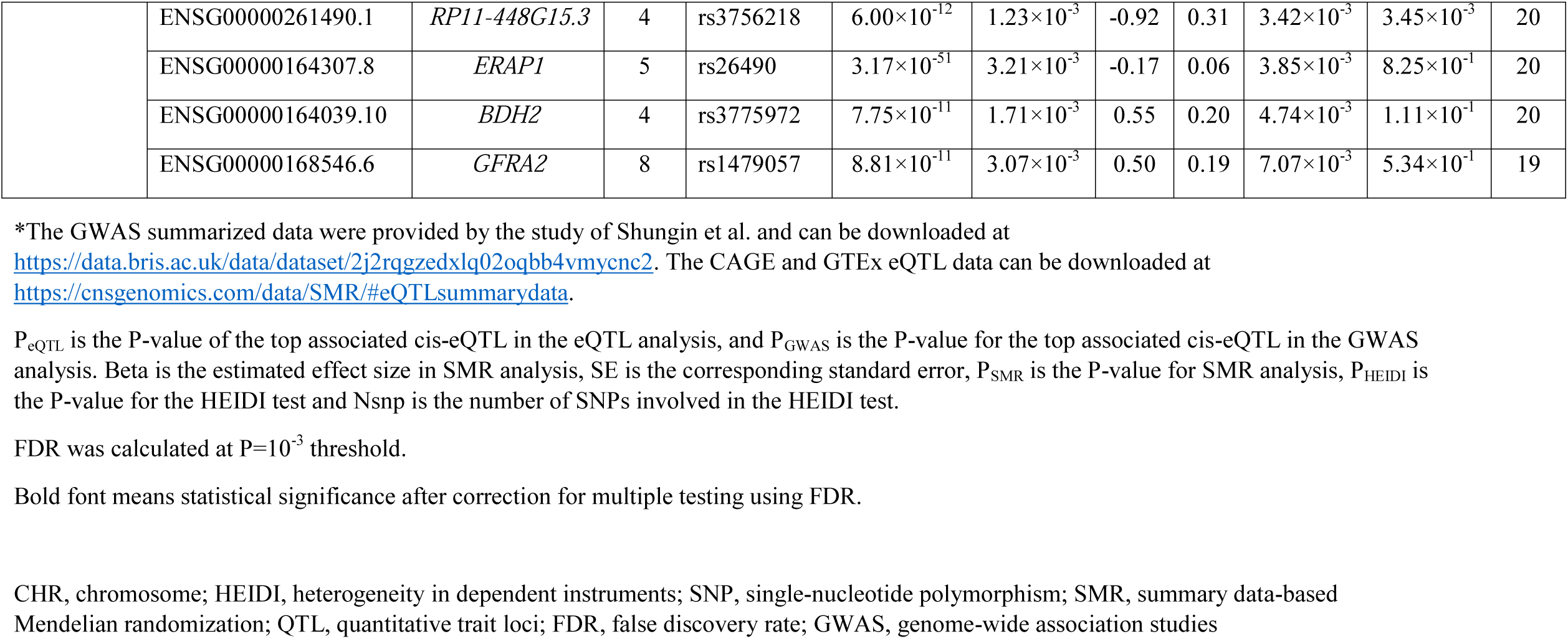
The top ten probes identified in the SMR analysis in participants of East Asian ancestry.

| eQTL data | Probe ID | Gene | CHR | Top SNP | P<sub>eQTL</sub> | P<sub>GWAS</sub> | Beta | SE | P<sub>SMR</sub> | P<sub>HEIDI</sub> | N<sub>SNP</sub> |
| --- | --- | --- | --- | --- | --- | --- | --- | --- | --- | --- | --- |
| **CAGE** | ILMN_1899903 | *BX093763* | 5 | rs984976 | $4.96 \times 10^{-39}$ | $1.27 \times 10^{-4}$ | 0.42 | 0.11 | **$2.33 \times 10^{-4}$** | $2.67 \times 10^{-1}$ | 20 |
| | ILMN_1734231 | *DDOST* | 1 | rs6893 | $8.13 \times 10^{-40}$ | $1.17 \times 10^{-3}$ | 0.27 | 0.09 | $1.62 \times 10^{-3}$ | $9.21 \times 10^{-2}$ | 20 |
| | ILMN_2154115 | *PSD4* | 2 | rs2241976 | $1.10 \times 10^{-77}$ | $1.53 \times 10^{-3}$ | 0.26 | 0.08 | $1.79 \times 10^{-3}$ | $2.64 \times 10^{-6}$ | 20 |
| | ILMN_2388155 | *CASP3* | 4 | rs11721363 | $1.33 \times 10^{-39}$ | $2.07 \times 10^{-3}$ | -0.88 | 0.29 | $2.72 \times 10^{-3}$ | $1.80 \times 10^{-6}$ | 20 |
| | ILMN_1764522 | *LMBR1* | 7 | rs73167977 | $3.88 \times 10^{-22}$ | $2.01 \times 10^{-3}$ | -0.74 | 0.25 | $3.26 \times 10^{-3}$ | $9.03 \times 10^{-4}$ | 20 |
| | ILMN_1656300 | *GFRA2* | 8 | rs1479056 | $2.14 \times 10^{-62}$ | $3.07 \times 10^{-3}$ | 0.30 | 0.10 | $3.55 \times 10^{-3}$ | $5.51 \times 10^{-1}$ | 20 |
| | ILMN_1791211 | *DOK2* | 8 | rs1479056 | $1.14 \times 10^{-47}$ | $3.07 \times 10^{-3}$ | 0.34 | 0.12 | $3.72 \times 10^{-3}$ | $1.65 \times 10^{-1}$ | 20 |
| | ILMN_1808661 | *TOMM5* | 9 | rs7018807 | $9.27 \times 10^{-61}$ | $3.58 \times 10^{-3}$ | -0.34 | 0.12 | $4.09 \times 10^{-3}$ | $4.20 \times 10^{-3}$ | 20 |
| | ILMN_1910292 | *BX094911* | 4 | rs3111820 | $3.02 \times 10^{-5}$ | $7.91 \times 10^{-5}$ | -1.46 | 0.51 | $4.13 \times 10^{-3}$ | $2.69 \times 10^{-1}$ | 4 |
| | ILMN_1805590 | *NAA38* | 7 | rs7799229 | $1.28 \times 10^{-67}$ | $3.93 \times 10^{-3}$ | 0.24 | 0.08 | $4.44 \times 10^{-3}$ | $9.13 \times 10^{-5}$ | 20 |
| **GTE<sub>x</sub>** | ENSG00000204792.2 | *AC104135.3* | 2 | rs12366 | $5.06 \times 10^{-108}$ | $6.39 \times 10^{-4}$ | 0.12 | 0.03 | **$7.46 \times 10^{-4}$** | $3.11 \times 10^{-1}$ | 20 |
| | ENSG00000204469.8 | *PRRC2A* | 6 | rs2075800 | $3.28 \times 10^{-8}$ | $6.54 \times 10^{-5}$ | 1.72 | 0.53 | $1.22 \times 10^{-3}$ | $8.73 \times 10^{-2}$ | 20 |
| | ENSG00000243753.1 | *HLA-L* | 6 | rs3094204 | $1.45 \times 10^{-17}$ | $6.48 \times 10^{-4}$ | -0.27 | 0.09 | $1.54 \times 10^{-3}$ | $1.97 \times 10^{-3}$ | 20 |
| | ENSG00000224769.1 | *AC069213.1* | 3 | rs6804822 | $3.13 \times 10^{-14}$ | $5.05 \times 10^{-4}$ | 0.42 | 0.13 | $1.56 \times 10^{-3}$ | $1.34 \times 10^{-3}$ | 20 |
| | ENSG00000125637.11 | *PSD4* | 2 | rs2241976 | $2.71 \times 10^{-66}$ | $1.53 \times 10^{-3}$ | 0.34 | 0.11 | $1.84 \times 10^{-3}$ | $1.96 \times 10^{-5}$ | 20 |
| | ENSG00000144791.5 | *LIMD1* | 3 | rs34448158 | $4.35 \times 10^{-11}$ | $5.24 \times 10^{-4}$ | 0.71 | 0.23 | $2.16 \times 10^{-3}$ | $5.00 \times 10^{-1}$ | 20 |
| | ENSG00000261490.1 | *RP11-448G15.3* | 4 | rs3756218 | $6.00 \times 10^{-12}$ | $1.23 \times 10^{-3}$ | -0.92 | 0.31 | $3.42 \times 10^{-3}$ | $3.45 \times 10^{-3}$ | 20 |
| | ENSG00000164307.8 | *ERAP1* | 5 | rs26490 | $3.17 \times 10^{-51}$ | $3.21 \times 10^{-3}$ | -0.17 | 0.06 | $3.85 \times 10^{-3}$ | $8.25 \times 10^{-1}$ | 20 |
| | ENSG00000164039.10 | *BDH2* | 4 | rs3775972 | $7.75 \times 10^{-11}$ | $1.71 \times 10^{-3}$ | 0.55 | 0.20 | $4.74 \times 10^{-3}$ | $1.11 \times 10^{-1}$ | 20 |
| | ENSG00000168546.6 | *GFRA2* | 8 | rs1479057 | $8.81 \times 10^{-11}$ | $3.07 \times 10^{-3}$ | 0.50 | 0.19 | $7.07 \times 10^{-3}$ | $5.34 \times 10^{-1}$ | 19 |

\*The GWAS summarized data were provided by the study of Shungin et al. and can be downloaded at https://data.bris.ac.uk/data/dataset/2j2rqgzexlq02oqbb4vmcnc2. The CAGE and GTEx eQTL data can be downloaded at https://cnsgenomics.com/data/SMR/#eQTLsummarydata.
$P_{\text{eQTL}}$ is the P-value of the top associated cis-eQTL in the eQTL analysis, and $P_{\text{GWAS}}$ is the P-value for the top associated cis-eQTL in the GWAS analysis. Beta is the estimated effect size in SMR analysis, SE is the corresponding standard error, $P_{\text{SMR}}$ is the P-value for SMR analysis, $P_{\text{HEIDI}}$ is the P-value for the HEIDI test and N<sub>snp</sub> is the number of SNPs involved in the HEIDI test.
FDR was calculated at $P=10^{-3}$ threshold.
Bold font means statistical significance after correction for multiple testing using FDR.
CHR, chromosome; HEIDI, heterogeneity in dependent instruments; SNP, single-nucleotide polymorphism; SMR, summary data-based Mendelian randomization; QTL, quantitative trait loci; FDR, false discovery rate; GWAS, genome-wide association studies

**Figure 3.**
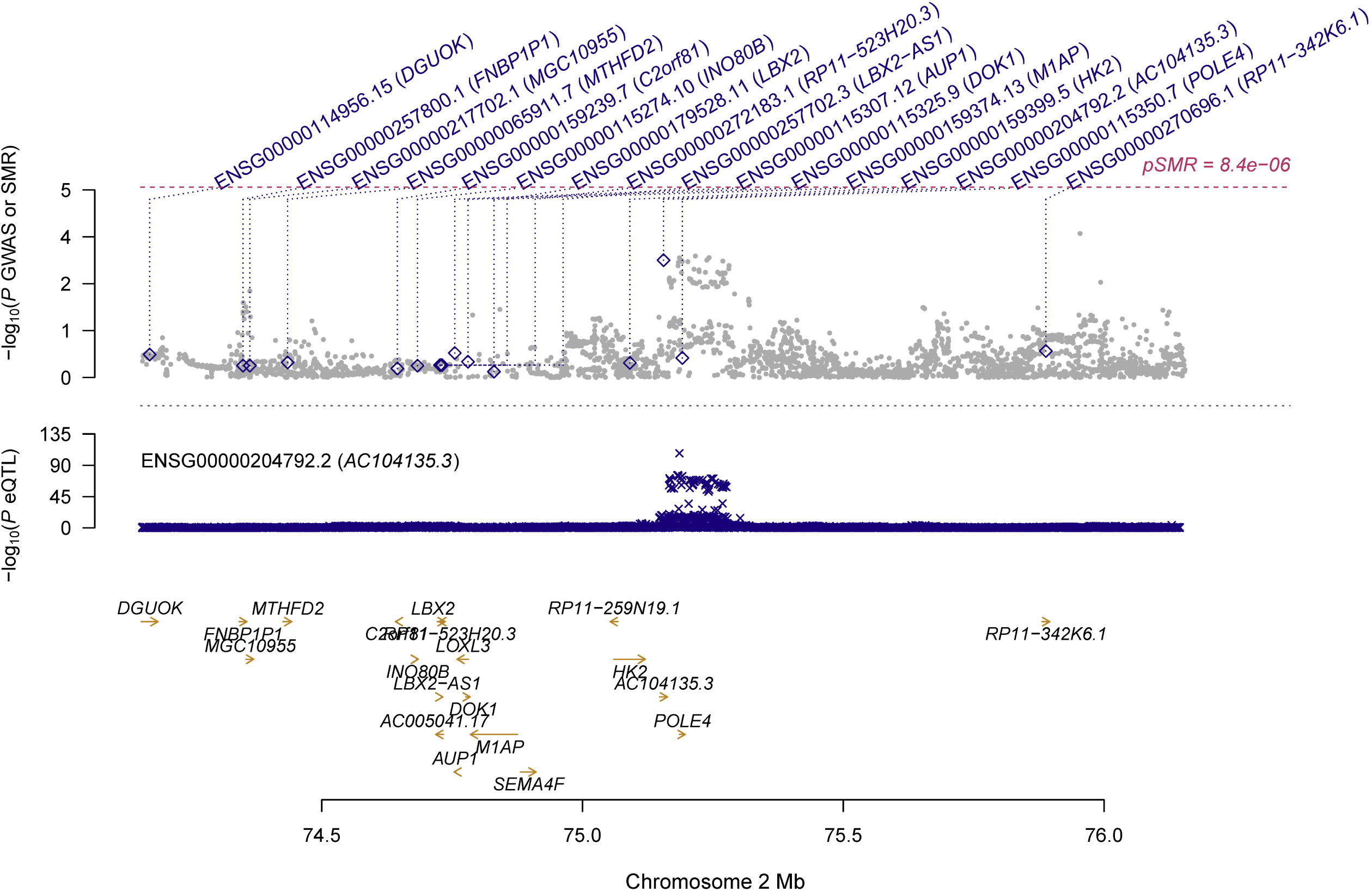
Prioritizing gene around *AC104135.3* in association with periodontitis in participants of East Asian ancestry. Top plot, grey dots represent the -log_10_(*P* values) for SNPs from the GWAS of periodontitis, and rhombuses represent the -log_10_(*P* values) for probes from the SMR test with solid rhombuses indicating that the probes pass HEIDI test and hollow rhombuses indicating that the probes do not pass the HEIDI test. Middle plot, eQTL results for ENSG00000204792.2 probe, tagging *AC104135.3*. Bottom plot, location of genes tagged by the probes. GWAS, genome-wide association studies; SMR, summary data-based Mendelian randomization; HEIDI, heterogeneity in dependent instruments; eQTL, expression quantitative trait loci

We found that two genes were among the top hits in both SMR analyses, including *PSD4* (CAGE eQTL: ILMN_2154115, P_SMR_=1.79×10^−3^; GTEx: ENSG00000125637.11, P_SMR_=1.84×10^−3^) and *GFRA2* (CAGE eQTL: ILMN_1656300, P_SMR_=3.55×10^−3^; GTEx: ENSG00000168546.6, P_SMR_=7.07×10^−3^). More information could be found in **Table S3-4**.

## Discussion

In the present study, we explored putative genes that showed pleiotropic/potentially causal association with periodontitis by integrating GWAS and eQTL data in the SMR analysis. We identified multiple genes, some of which represented novel genes, that might be involved in the pathogenesis of periodontitis in participants of European ancestry and participants of East Asian ancestry. Our findings provided helpful leads to a better understanding of the mechanisms underlying periodontitis and suggested potential therapeutic targets for the treatment of periodontitis.

A recent study investigated molecular biomarker candidates and biological pathways of chronic periodontitis using pooled datasets in the Gene Expression Omnibus (GEO) database, and identified 123 common differently expressed genes (DEGs), including 81 upregulated genes and 42 downregulated genes^32^. Several of the identified genes were also among the top hits in our SMR analysis. For example, the gene *NSG1* (Neuronal Vesicle Trafficking Associated 1) was found to be downregulated in persons with chronic periodontitis. It also showed significant pleiotropic association with periodontitis in our study of participants of European ancestry (**Table 2**). *NSG1*, also known as *D4S234E* or *NEEP21*, is located on 4p16.3 in human and is a member of the neuron-specific gene family. It is the most important in regulating receptor recycling and synaptic transmission^33^. p53, an important tumor suppressor gene, binds to the promoter region of *NSG1* and regulates its expression in response to DNA damage. Inhibition of *NSG1* expression suppressed apoptosis^34^. The exact role of *NSG1* in the pathogenesis of periodontitis is unclear and warrants further research.

Another research integrating GWAS and eQTL data identified 10 genes whose expression might influence periodontitis^35^. Of them, the gene *S100A12* (S calcium-binding protein A12) also appeared among the top hits in participants of European ancestry in the SMR analysis using CAGE and GTEx eQTL data (**Table 2**). *S100A12*, located on 1q21.3, is a member of the S100 family of EF-hand calcium-binding proteins^36^. Previous studies indicated that it played a prominent role in the regulation of inflammatory processes and immune response^37^. It was reported that the levels of *S100A12* were higher in participants with high periodontal inflammatory burden and were associated with the percentage of bleeding on probing^38^. In gingival crevicular fluid and serum, the levels of *S100A12* increased with the inflammation of periodontium^39^. These findings, together with ours, demonstrated the important role of *S100A12* in influencing periodontitis and highlighted the potential of this gene as a promising target for the prevention and treatment of periodontitis.

Our study was different from the previous study integrating GWAS and eQTL data^35^. We used the GWAS summarized data for both European and East Asian ancestries, while the previous research only analyzed GWAS data of European ancestry. Similarly, we used both CAGE and GTEx eQTL data, while the previous research used only GTEx data. Moreover, we undertook a SMR analytic framework which focused on exploring genes showing pleiotropic association/potentially causal association with periodontitis while the previous research adopted a Sherlock approach which is a Bayesian statistical framework aiming to identify genes whose expression was associated with periodontitis susceptibility^40^.

Our study was also very different from another MR research on periodontitis^29^. Although both studies aimed to explore potentially causal factors for periodontitis, the previous research focused examining the causal role of total adiposity in the pathogenesis of periodontitis, while our study attempted to identify genes that were pleiotropically/potentially causally associated with periodontitis. The analytic approaches were also different: the previous research used genetic risk scores as the instrumental variables calculated from three genes (*FTO, MC4R* and *TMEM18*) by summing the number of BMI increasing alleles, while in our study, we used all the genetic variants from GWAS summarized data as the potential instrumental variables.

Our study has some limitations. The number of probes used in our SMR analysis was limited, especially in the SMR analysis of participants of East Asian ancestry. As a result, we may have missed some genes which played important roles in the pathogenesis of periodontitis. The HEIDI test was significant for some of the identified genes (**Table 2-3**). Therefore, we could not rule out the possibility of horizontal pleiotropy, i.e., the identified association might be due to two distinct genetic variants in high linkage disequilibrium with each other. In addition, we only performed SMR analysis for participants of European and East Asian ancestries, and our findings might not be generalized to other populations. More studies are needed to validate our findings in independent populations. Due to the exploratory nature of study, we adopted correction for multiple testing to reduce false positive rate; however, we may have missed important SNPs or genes. Finally, we could not quantify the changes in gene expression in subjects with periodontitis in comparison with the control due to the unavailability of individual eQTL data.

## Conclusions

In conclusion, our SMR analysis revealed multiple genes that were potentially pleiotropically associated with periodontitis. More studies are needed to explore the underlying physiological mechanisms in the etiology of periodontitis.

## Data Availability

All data generated or analyzed during this study are included in this published article and its supplementary information files.

## Acknowledgements

The study was supported by NIH/NIA grants P30AG10161, R01AG15819, R01AG17917, R01AG36042, U01AG61356 and 1RF1AG064312-01. Di Liu was supported by China Scholarship Council (CSC 201908110339). The authors confirmed that all authors have reviewed the contents of the article being submitted, approved its contents, and validated the accuracy of the data.

## Author contribution

DL, JY and GZ designed the study. FW, DL, BF and WL analyzed data and performed data interpretation. FW, DL, YZ and JY wrote the initial draft and BF, WL, and GZ contributed to writing subsequent versions of the manuscript. All authors reviewed the study findings and read and approved the final version before submission.

## Figure legends

**Figure S1.**
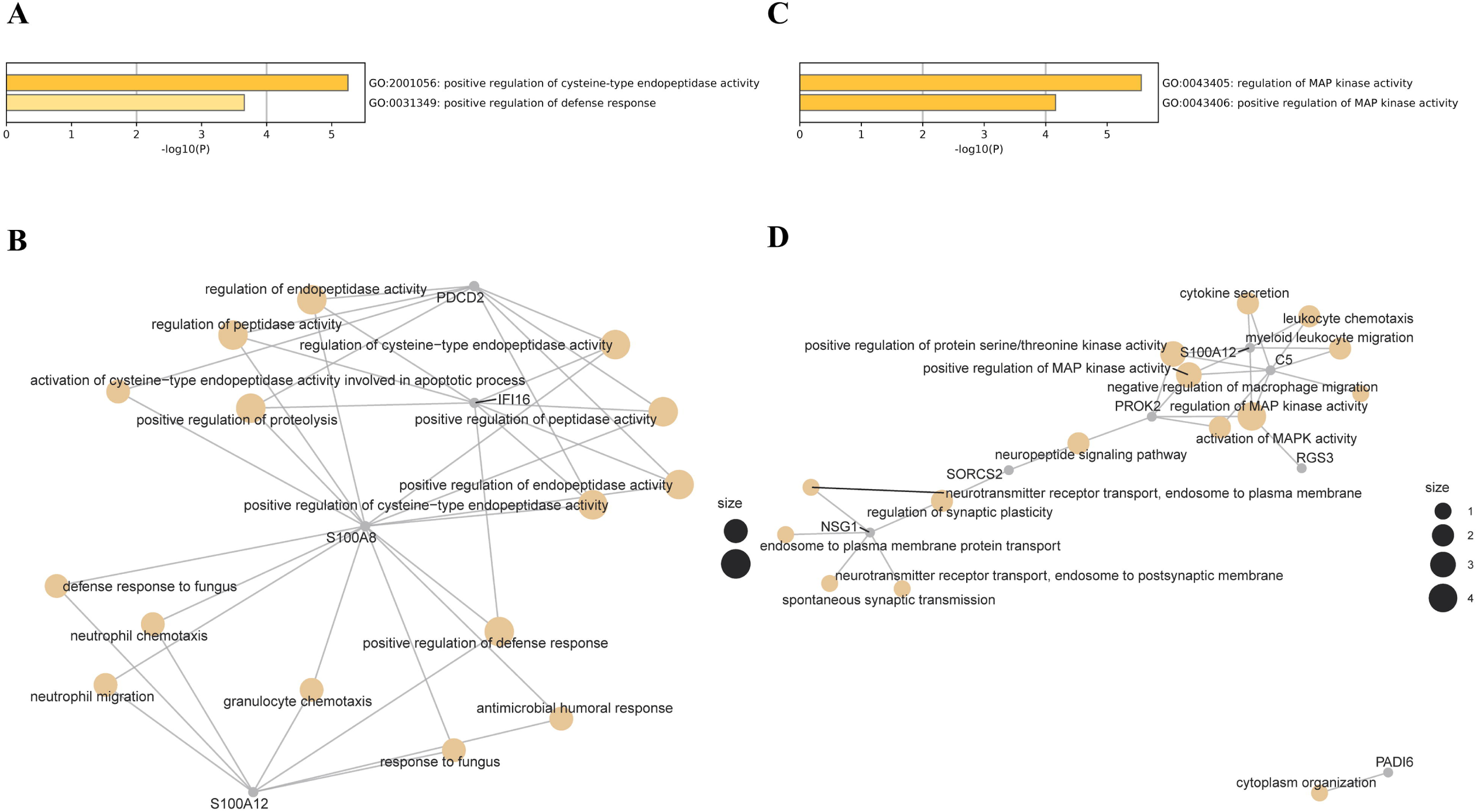
Functional enrichment and gene concept network analysis in participants of European ancestry. A) Enriched GO terms based on genes identified using CAGE eQTL data; B) Concept network analysis of the genes identified using CAGE eQTL data; C) Enriched GO terms based on genes identified using GTEx eQTL data; and D) Concept network analysis of the genes identified using GTEx eQTL data. GO, gene ontology; eQTL, expression quantitative trait loci

**Figure S2.**
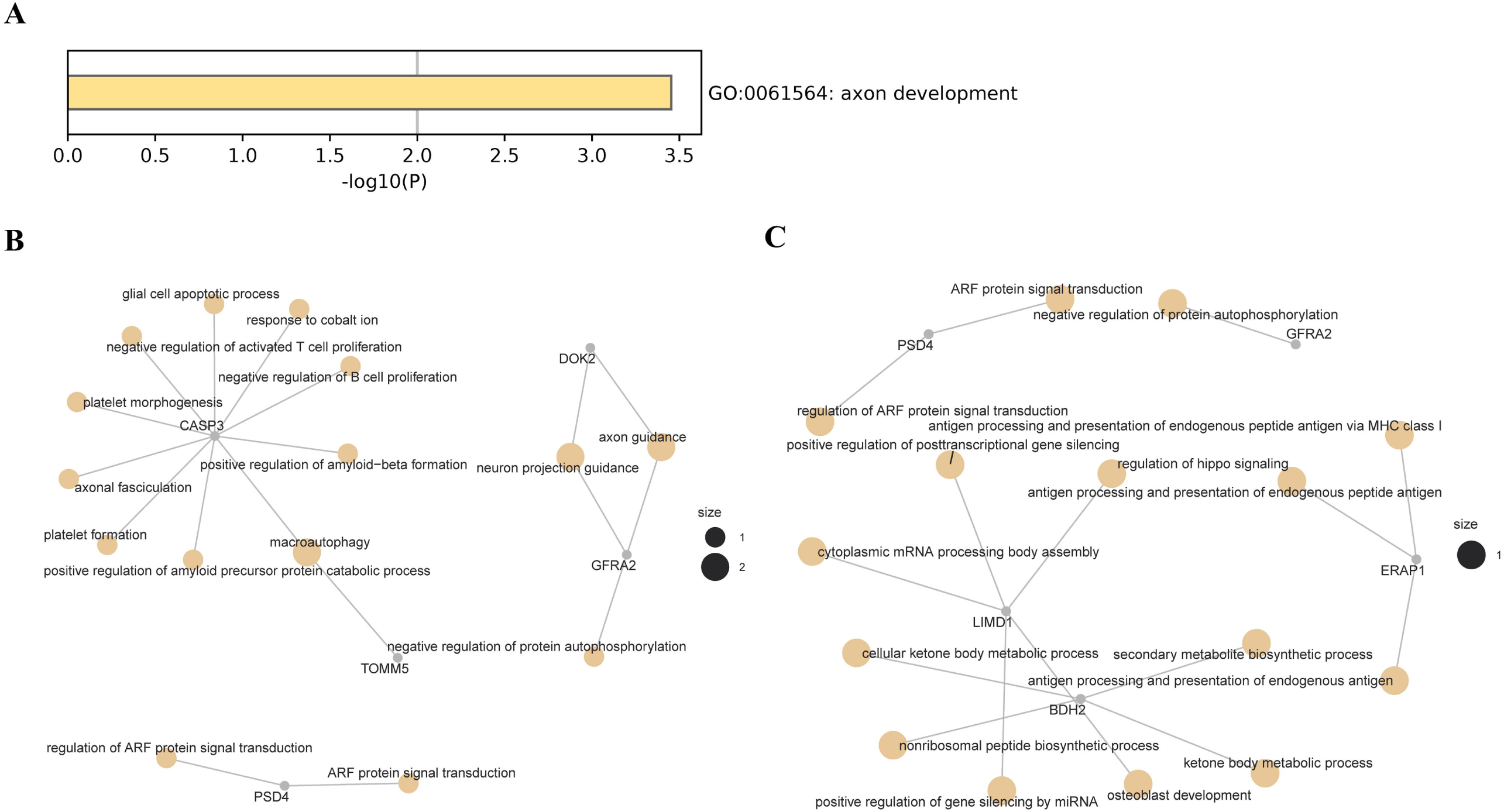
Functional enrichment and gene concept network analysis in participants of East Asian ancestry. A) Enriched GO terms based on genes identified using CAGE eQTL data; B) Enriched GO terms based on genes identified using GTEx eQTL data; and C) Concept network analysis of the genes identified using GTEx eQTL data. GO, gene ontology; eQTL, expression quantitative trait loci

